# Protein and Microbial Biomarkers in Sputum Discern Acute and Latent Tuberculosis in Investigation of Pastoral Ethiopian Cohort

**DOI:** 10.1101/2020.09.02.20182097

**Authors:** Milkessa HaileMariam, Yanbao Yu, Harinder Singh, Takele Teklu, Biniam Wondale, Adana Worku, Aboma Zewde, Stephanie Monaud, Tamara Tsitrin, Mengistu Legesse, Gobena Ameni, Rembert Pieper

## Abstract

Differential diagnosis of tuberculosis (TB) and latent TB infection (LTBI) remains a public health priority in high TB burden countries. Pulmonary TB is diagnosed by sputum smear microscopy, chest X-rays, and PCR tests for distinct *Mycobacterium tuberculosis* (Mtb) genes. Clinical tests to diagnose LTBI rely on immune cell stimulation in blood plasma with TB-specific antigens followed by measurements of interferon-γ concentrations. The latter is an important cytokine for cellular immune responses against Mtb in infected lung tissue. Sputum smear microscopy and chest X-rays are not sufficiently sensitive while both PCR and interferon-γ release assays are expensive. Alternative biomarkers useful for developing diagnostic tests to discern TB disease states are desirable. This study’s objective was to discover biomarkers in sputum, assessing the proteomes and microbiomes of 74 TB patients, 46 individuals with LTBI, and 51 negative community controls (NCC). Study participants were from the South Omo province, a pastoral region in southern Ethiopia. A total of 161 and 115 samples were used to determine the 16S rRNA sequence-based bacterial taxonomies and proteomic profiles, respectively. Sputum microbiota did not reveal statistically significant differences in α-diversity comparing the three groups. The genus *Mycobacterium*, representing Mtb, was only identified for the TB group. The latter featured reduced abundance of the genus *Rothia* in comparison to the LTBI and NCC groups. *Rothia* is a human respiratory tract commensal and may be sensitive to the inflammatory milieu caused by TB infection. Proteomic data strongly supported innate immune responses against Mtb in subjects with pulmonary TB. Ferritin, an iron storage protein released by damaged host cells, was markedly increased in abundance in TB sputum compared to the LTBI and NCC groups, along with α-1-acid glycoproteins ORM1 and ORM2. These proteins are acute phase reactants and inhibit excessive neutrophil activation. Proteomic data also supported effector roles of neutrophils in the anti-Mtb response which was not observed for LTBI cases. Less abundant in sputum of the LTBI group versus the NCC group were two immunomodulatory proteins, mitochondrial TSPO and the extracellular ribonuclease T2. If validated, these proteins are of interest as diagnostic biomarkers for LTBI.

## 1 INTRODUCTION

Diagnosis of active tuberculosis (TB) and latent TB infection (LTBI) is important to control the spread of *Mycobacterium tuberculosis* (Mtb), a persistent and increasingly multi-antibiotic drug resistant (MDR) pathogen. TB remains an urgent public health issue in thirty high disease burden countries [1]. Eight to twelve million Mtb infection cases per year advance to stages with clinical symptoms; 90% of all cases do not cause systems [2]. The primary manifestation is pulmonary TB (PTB). Extrapulmonary TB which affects other organs is less prevalent [3]. Severity of TB is influenced by intrinsic (genetic) factors [4-7] and extrinsic (environmental) factors such as nutrition and immunological status of the host. Helminth infections, and particularly HIV/AIDS are comorbidities prevalent in African countries and increase risk of progression to symptomatic TB [3, 8]. Less well investigated has been the role of distinct oral and alveolar niche microbiomes as modulators of disease outcome [9, 10]. In PTB, the focus of this study, transmission occurs through inhalation of aerosolized bacteria in droplets.

Certain Mtb strains are more transmissible than others [11], but the reasons for differences in the severity of patient outcomes appear to be linked to the immune response. Mtb invades and manipulates the function of alveolar macrophages, resulting in the formation of granulomas that consist of differentiated epithelioid and multinucleated macrophages, dendritic cells, CD4 and CD8 T-cells as well as B-cells at the infection site [12]. Key cytokines for the immune responses against Mtb in the lungs are interferon-γ (IFN-γ) and tumor necrosis factor-α (TNF-α). While IFN-γ is mostly produced by T-cells, TNF-α is released by T-cells and macrophages [12]. Neutrophils are recruited to the infection site and contribute to local inflammation and pathogen containment. In the chronic phase of TB, a balance between Th1 and Th17 responses controls bacterial growth and limits the immunopathology [13]. Bacterial pathogen cells largely reside in the macrophages of granulomas. The latter act as barriers to bacterial spread to other regions of the lungs. Individuals with delayed-type hypersensitivity responses to TB antigens such as ESAT-6 and CFP10 and lack of symptoms are deemed to be latently infected. While not contagious [12], they are susceptible to disease activation at a later timepoint. IFN-γ release assays (IGRAs) that relies on the stimulation of blood plasma immune cells with TB antigens followed by quantitative measurements of secreted IFN-γ are the current gold standard of LTBI diagnosis [14]. Using cytokine antibody arrays, we surveyed the blood plasma of TB, LTBI, and negative community controls (NCC) from the cohort under study here and identified RANTES as a potential plasma biomarker of LTBI. The chemokine was differentially abundant with statistical significance (LTBI vs TB), with and without prior TB antigen stimulation [15].

Sputum is a great source of TB biomarker discovery due to the low health risk associated with sampling and proximity to infection sites. Widely used, but slow or not sufficiently sensitive diagnostic TB tests are acid-fast bacilli smear microscopy and isolation of Mtb strains from sputum culture, respectively [16]. The PCR-based test Xpert MTB/RIF assay (Cepheid, USA) has higher sensitivity for Mtb detection in smear-positive than in smear-negative patients and also detects rifampicin resistance allowing assessment of the antibiotic treatment options [16]. Sputum is also a biomarker source to measure inflammation of the lungs derived from other diseases, such as cystic fibrosis [17]. Such biomarkers may not be specific for a respiratory pathogen, irritant or intrinsic factor. Quantitative changes of metabolites and proteins have been evaluated in clinical samples for the diagnosis, latency and progression of TB or treatment responses. Miranda et al. detected sustained levels of ferritin and C-reactive protein (CRP) in patients who remained Mtb culture-positive during antibiotic treatment of TB in a Brazilian cohort. These biomarkers indicated persistence of lung inflammation as compared to those who became culture-negative [18]. Gopal et al. identified calcium-mobilizing calprotectins as mediators of neutrophil-linked inflammation in granuloma-positive PTB patients and suggested that silencing their activities may attenuate patient symptoms [19]. Goletti et al. highlighted the need of developing predictive biomarkers as surrogate endpoints in clinical trials for new investigational TB drugs [20]. Using highly parallel “omics” analysis, a neutrophil-driven blood transcriptional signature induced by IFN-γ was identified for active TB [21], consistent with neutrophil infiltration of the infected lungs. Jiang et al. demonstrated reduced CD27 expression in TB antigen-targeting CD4 T cells during persistent infection and commented on the role of this T-cell surface biomarker in chronic TB diagnosis [22]. Of particular interest are biomarkers that discern LTBI from healthy controls since there is a risk of pathogen reactivation. One study revealed proteomic fingerprints in TB antigen-stimulated plasma of patients describing greater than 82% specificity and 89% sensitivity distinguishing TB from LTBI [23]. A proteomic approach was reported to discern LTBI from healthy controls at greater than 85% specificity and sensitivity using non-stimulated plasma samples [24]. Protein biomarkers suggesting protective effects of vaccines are of high clinical interest. Gopal et al. found IL-17 to be involved in protective immunity against a virulent Mtb strain. A role of IL-17 in driving Th1 cell responses upon BCG vaccination was suggested [25, 26].

The impact of oral and respiratory tract microbiomes on human health are in an exploratory phase and difficult to study due to the enormous complexity of microbiome-host relationships [27]. Distinct respiratory tract-colonizing bacterial taxa may correlate with, cause susceptibility to, or mediate protection from lung diseases, including TB. Causal relationships were demonstrated for asthma development in infants, for example [28]. Studies of oral and respiratory tract microbiomes in the context of TB patients have been limited to date, as reviewed by Hong et al. [29] and Naidoo et al. [30]. Our objectives were to analyze sputum microbiomes and proteomes for an interesting multi-ethnic cohort of Ethiopian pastoralists and to interpret the data to gain new insights into the host-pathogen-microbiome crosstalk and to identify disease outcome-related biomarkers.

## 2 MATERIALS AND METHODS

### 2.1 Human Subjects and TB Diagnostic Approaches

Human subject recruitment occurred in a multi-ethnic southern Ethiopian region (South Omo province). Covering eight districts, a study named “Systems Biology for Molecular Analysis of Tuberculosis in Ethiopia” resulted in informed consent of ~ 2,100 individuals aged 15 years or older at local or regional clinics. Some individuals had symptoms consistent with TB. Negative community controls (NCC) often were household contacts of those suspected to be infected with TB. Nearly 1,200 sputum and blood samples were collected. Of those, nearly 13% were positive using acid-fast bacilli (AFB) smear microscopy or by isolating Mycobacterium tuberculosis complex (MTBC) strains on Lowenstein Jensen medium [31]; 50.5% were positive for LTBI employing IFN-γ assays [32]. Subjects positive for PTB were offered DOTS treatment [33] at a clinic close to their residences. Information on geographic and socio-economic characteristics, ethnic affiliations, exclusion from participation, and other medical data were reported recently [31, 32]. The Institutional Review Board at Aklilu Lemma Institute of Pathobiology, Addis Ababa University (ALIPB-AAU) under ref. no. ALIPB/IRB/22-B/2012/13, the National Research Ethics Committee of Ethiopia (ref no. 3.10/785/07), and the J. Craig Venter Institute (JCVI) IRB (ref. no. 2014-200) approved the human subject protocol. The LTBI sub-group was later distinguished from NCC subjects based on data from QuantiFERON-TB Gold In-Tube test (GFT-GIT) assays. We set the positivity threshold for LTBI at the recommended ≥ 0.35 IU/ml [32].

### 2.2 Sputum Sample Collection and Processing

Study participants were instructed to cough up sputum. It was collected in pre-labeled cups. Where possible, subjects completed three cycles of sputum expectoration using 3-5% hypertonic sodium chloride. The single-timepoint specimens were combined and processed as described previously [34]. Briefly, collected sputum aliquots for proteome and microbiome analysis were disinfected with SDS buffer (1% SDS, 10 mM Na-EDTA, 50 mM DTT, 0.03% Tween-20, 50 mM Tris/Tris-HCl, pH 8.0) and centrifuged at 16, 000 x g to recover the supernatants. SDS-denatured and heat-treated lysates (85°C) were shipped to the site of “omics” analyses (JCVI) and stored long-term at −80°C prior to analyte extraction. Protein samples were run in SDS-PAGE gels (4-12%T) to estimate total protein concentrations. Aliquots of 150 μg protein extract were subjected to S-Trap Ultra-Fast sample preparation [34], digesting proteins with trypsin at a 50:1 mass ratio. Peptides were desalted prior to LC-MS/MS analysis using the spinnable StageTip protocol [35]. rDNA extraction and analysis are described below.

### 2.3 Shotgun proteomics

An Ultimate 3000-nano liquid chromatography (LC) system coupled to a Q-Exactive mass spectrometer (both units from Thermo Scientific) was used for LC-MS/MS analysis. The workflow and data acquisition methods have been described comprehensively [36]. Briefly, peptide digestion products of approximately 10 μg were separated over a 150 min gradient from 2% to 80% acetonitrile (120 min to 35%, 10 min to 80%), with 0.1% formic acid in buffers A and B. The flow rate using an in-house packed column (75 µm x 15 cm, 3.0 µm ReproSil-Pur C_18_-AQ media) was 200 nl/min. MS survey scans were acquired at a resolution of 70,000 over a mass range of m/z 350-1,800. During each cycle in a data-dependent acquisition mode, the ten most intense ions were subjected to high-energy collisional dissociation (HCD) applying a normalized collision energy of 27%. MS/MS scans were performed at a resolution of 17,500. Two technical replicate LC-MS/MS runs were performed per sample, and the MS data were combined in the MaxQuant analysis process.

### 2.4 Identification and Quantitation of Proteins

The MS raw data were processed using the Proteome Discoverer platform (version 1.4, Thermo Scientific) and the Sequest HT algorithm. The database contained protein sequences from the Mtb strain ATCC 25618/H37Rv (7,955 entries) and human proteins (20,195 sequence entries; reviewed sequences only; version 2015_06). The search parameters included two missed tryptic cleavages, oxidation (M), protein N-terminal acetylation and deamidation (N, Q) as variable modifications, and carbamidomethylation (C) as a fixed modification. Minimum peptide length was seven amino acids. The MS and MS/MS ion tolerances were set at 10 ppm and 0.02 Da, respectively. The false discovery rate (FDR) was estimated employing the integrated Percolator tool. Only protein hits identified with a 1% FDR threshold were accepted. For protein quantification, the MaxQuant and Andromeda software suite (version 1.4.2.0) was used, accepting most default settings provided in the software tool [37]. Label-free quantification (LFQ) generates relative protein abundance data from integrated MS^1^ peak areas of the high-resolution MS scans [38]. Only proteins quantified by at least one unique peptide were used for analysis. LFQ values were log (base 2) transformed, and then imputed with respect to missing values. Clustering and correlation analyses were performed using functions embedded in the Perseus (version 1.5.0.15) software. The LC-MS/MS data were deposited in ProteomeXchange via the PRIDE partner repository under the dataset identifier is PXD012412.

### 2.5 16S rDNA Sequencing

Aliquots of 300 μl of the SDS-denatured sputum lysates were subjected to phenol-chloroform extraction to isolate total DNA. Ethanol-precipitated enriched DNA extracts were subjected to PCR amplification using 515F and 806R forward and reverse primers, respectively, to amplify the V4 region of microbial DNA in the extracts. The PCR amplification method was previously described [39]. A two-step amplification was performed to reduce the risk of non-specific binding when using adapters/sequencing primers of more than 100 base pairs (bp). For 167 individual specimens, sufficient quantities of the V4 sequence region (254 bp) were amplified as visualized by ethidium bromide staining in agarose gels. Samples for DNA library preparation were obtained by excising bands of approximately 300 bp and normalizing the DNA quantity per sample by quantifying with Quanti-iTTM PicoGreen® (Life Technologies, USA). Amplicons were pooled at 100 ng each. A positive control (Escherichia coli DNA extract) was subjected to the same amplification and purification protocol. A standard Illumina sequencing-based library preparation and sequencing protocol (MiSeq Reagent Kit v3, 600 cycles) was used as described [39]. The library dilution had 15% PhiX as an internal control at a 4 pM concentration.

### 2.6 Processing and Filtering of 16S Sequence Reads

We generated operational taxonomic units (OTUs) de novo from raw Illumina 16S rDNA sequence reads using the UPARSE pipeline (Edgar 2013). Methods for trimming of the adapter sequences, barcodes and primers, the elimination of sequences of low quality, the de-replication step and sequence read abundance determinations were analogous to those applied previously [40]. Chimera filtering of the sequences occurred during the clustering step. We used the Wang classifier, bootstrapping using 100 iterations and mothur to report full taxonomies for only those sequences where 80 or more of the 100 iterations were the same (cutoff=80). The taxonomies were assigned to OTUs using mothur [41] with the version 123 of the SILVA 16S ribosomal RNA database as the reference [42]. From the tables of OTUs with corresponding taxonomy assignments, we removed likely non-informative OTUs (rare OTUs and taxa strongly affected by MiSeq sequencing errors). Unbiased metadata-independent filtering was applied at each taxonomy level by eliminating features that did not pass the selected criteria (< 2000 reads and OTUs present in less than 10 samples), as described previously [40].

### 2.7 Identification of Phylogenetic Groups for Gut and Oral Microbiota

The phyloseq package version 1.16.2 in R package version 3.2.3 was used for the microbiome census data analysis [43]. The plot_richness function was used to create a plot of alpha diversity index estimates for each sample. The differences in microbial richness (α-diversity) were evaluated using Wilcoxon t-test. The ordination analysis was performed using the non-metric multi-dimensional scaling (NMDS) with the Bray-Curtis dissimilarity matrix [44]. The data output was used for the generation of a heatmap using the plot_heatmap function including a side bar where clinical variables associated with each sample were assigned to look for specific associations [43].

### 2.8 Statistical Analyses of Microbiome and Proteomic Data

To detect differential abundances in microbiota at a genus or species level the DESeq2 package version 1.12.3 in R was used. Phyloseq data are converted into a DESeq2 object using the function phyloseq_to_deseq2 function. DESeq2 [45] is a method for the differential analysis of count data that uses shrinkage estimation for dispersions and fold changes to improve both stability and interpretability of the estimates. The DESeq2 test uses a negative binomial model rather than simple proportion-based normalization or rarefaction to control for different sequencing depths, which may increase the power and also lower the false positive detection rate [46]. Default options of DESeq2 were used for multiple testing adjustment applying the Benjamini-Hochberg method [47]. To detect differential abundances in proteomic datasets, we used the Welch t-test, an unequal variance two-sample location test embedded in the Perseus software tool. The P-value significance threshold was set at ≤ 0.01. To identify enriched biological pathways from differentially abundant proteins (TB vs LTBI), we employed GO term analysis (http://geneontology.org/). To determine the enriched expression of proteins in tissues or anatomical locations, we reviewed relevant information in the Protein Atlas database (https://proteinatlas.org/).

### 2.9 Western Blotting

Approximately 10 μl of the sputum lysates were run in 4-12%T SDS-PAGE gels, electroblotted onto PVDF membranes (1.5h at 150 V), and incubated with antibody dilutions separated by intermittent PBS/0.1% Tween-20 wash steps as reported earlier [48]. A primary anti-human α-1-acid glycoprotein (α1AGP) polyclonal IgG fraction antibody developed in rabbit (Sigma-Aldrich; A0534) was used at a 1:2,000 dilution (overnight at 4°C). A secondary goat anti-rabbit IgG-HRP conjugate (Sta. Cruz Biotech; sc-2004) was used at a 1:10,000 dilution (2 h at 20°C). HRP-catalyzed color development with 3,3’-diaminobenzidine (DAB) required circa 5 minutes of incubation at 20°C.

## 3 RESULTS

### 3.1 Human subjects

Inhabitants of the South Omo province in southern Ethiopia are highly diverse with respect to ethnicities, cultures, and languages. Many of its people have a traditional lifestyle as cattle herders or in subsistence agriculture with little access to medical care. A subset of the nearly 2,000 participants in a molecular epidemiological study of TB [32] were used here to analyze sputum proteomes and microbiomes. The major ethnic groups were Hamar, Daasanech, Bena, Tsemay, Selamago, Maale, Ari, and Nyangatom. The prevalence of TB and LTBI were of interest given the diverse inhabitants, lack of urbanization, and increases in tourism. We reported on the MTBC lineage diversity for TB-positive subjects, the low prevalence of isolates resistant to first-line antibiotic drugs, and the high prevalence of LTBI [15, 31, 32]. Human subject ages ranged from 12 to 70 (mean = 38.5) and 46% were female. We analyzed 115 sputum samples using proteomics, and 161 samples using microbiome surveys. The datasets were categorized into the PTB, LTBI and NCC groups as specified in the Methods and Materials section and as denoted in Supplementary File S1.

### 3.2 Sputum proteome

Collapsing identifications regardless of disease group, 2,039 and 207 non-redundant human and Mtb proteins were obtained, respectively. The numbers pertain to those proteins represented by at least 2 unique peptides (Supplementary File S2). Mycobacterial proteins are not discussed in this report. LC-MS/MS technical replicates had higher correlation values for protein abundance than datasets where different biological samples were compared (R-values of 0.93-0.98 vs 0.57-0.76, as shown in Supplementary File S3). This was indicative of good quantitative accuracy achieved by the proteomic workflow and computational analysis. To assess the protein contributions from upper respiratory tract (saliva) and lower respiratory tract (expectorated sputum), we compared the data with three other studies, two of those for saliva [49-51]. The Venn diagram in Fig. 1 displays protein identification (ID) overlaps among all datasets. As expected, protein ID overlaps of our study and the one by Cao et al. on a sputum proteome [49] was highest (~87%). We conclude that sputum proteomes contain proteins derived from both upper and lower respiratory tract origin. Examples of proteins present in our data that have been reported to be enriched in saliva and sputum are the basic salivary proline-rich protein 3 (PRB3) [52] and the pulmonary surfactant-associated protein A1 (SFTPA1) [53], respectively.

**Figure 1.**
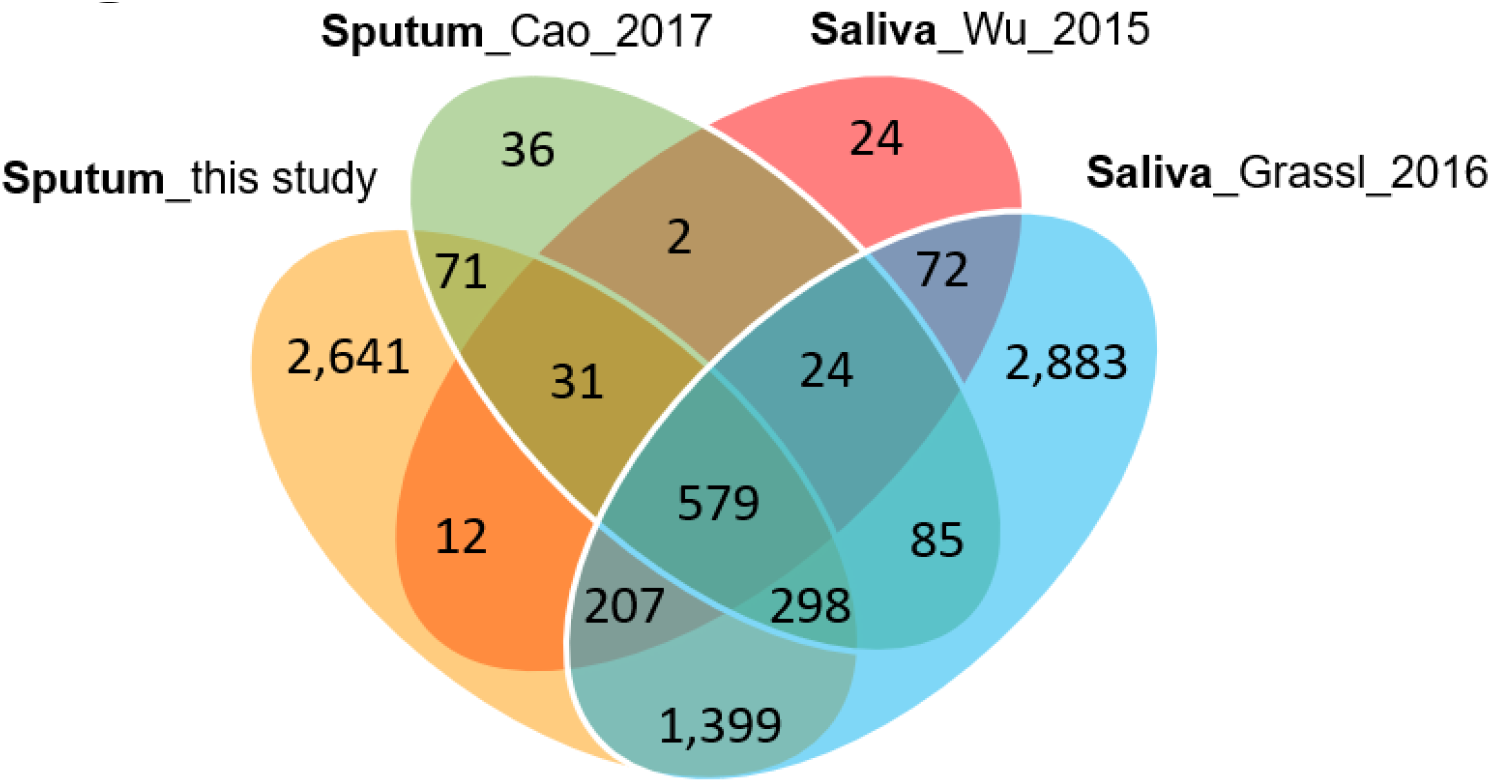
Venn diagram with protein identifications derived from two studies of saliva proteomes and two studies of sputum proteomes. The Cao study included analysis of samples associated with asthma. The Wu study included samples associated with squamous epithelial cell carcinoma of the oral cavity. In all studies, at least 800 proteins were identified.

### 3.3 Sputum proteomics reveals neutrophil infiltration and acute phase responses in the respiratory tract of PTB subjects

We selected datasets with at least 150 protein IDs per sample and protein IDs detected in at least nine subjects; 75 datasets were retained for quantification and clustering analyses, each with 432 distinct proteins (Supplementary File S2). Pearson Correlation hierarchical clustering (HCPC) was used to assess whether gender-specific differential analyses were warranted. While HCPC did not show significant gender-specific clustering, clusters were observed based on presence of infection (PTB) (Supplementary File S4). Next, the (gender-integrated) data were submitted to a Principal Component Analysis (PCA). PTB datasets largely clustered separately, although some overlap with the LTBI and NCC groups was observed (Fig. 2). Lack of separation between LTBI and NCC datasets in the PCA suggested that latency of TB does not strongly influence the sputum proteome compared to absence of infection. Differential sputum proteome analysis using unequal variance Welsh t-tests [54] resulted in 103 proteins with≥2-fold changes (Benjamini-Hochberg method corrected P-values < 0.01; PTB versus LTBI data). Twelve proteins are listed in Table 1. All 103 proteins are displayed in the Volcano Plot of Fig. 3. Detailed data are presented in Supplementary File S5. Protein functions and cell-specific expression were evidence of immune responses towards Mtb and inflammation in the PTB group alone: 16 and 14 of the 47 proteins increased in the PTB group (vs LTBI) are acute phase reactants or are highly expressed in leukocytes, respectively. The Protein Atlas tissue expression profiles supported that the 14 proteins were mostly released from neutrophils. All of them are expressed in neutrophils, and four of them had consensus data for unique presence in neutrophil granules or membranes (Supplementary File S5). Other differentially abundant proteins have oxidative stress response functions (superoxide dismutase 2, apolipoprotein D, catalase, and mitochondrial glutathione reductase). GO term biological process enrichment (Fig. 4) and protein network analysis in Cytoscape (Fig. 5) were consistent with leukocyte (primarily neutrophil)-mediated immune and acute phase response pathways. Neutrophils generate inflammation and kill pathogens using various mechanisms, while acute phase and oxidative stress responses limit the consequences of host cell collateral damage. Of the 56 proteins decreased in the PTB datasets in comparison to the LTBI datasets half are expressed, often distinctly enriched, in esophageal and oral mucosal tissues (Table 1; Supplementary File S5). Four of the proteins are secreted by salivary glands. Mucosal cells express and release specialized proteins important to form a barrier to the external environment. Protein network analysis and enriched GO term categories supported that relative quantitative increases of the 56 proteins (LTBI vs PTB) are associated with processes reflecting normal physiology of the respiratory tract squamous epithelium such as keratinocyte differentiation, keratinization, peptide cross-linking, stress responses, and epidermal development (Figs. 4 and 5). Given that inflammatory cells and proteins are released into sputum during anti-Mtb responses, those that represent normal squamous epithelial secretion and cell shedding are reduced in abundance, relatively to the LTBI group.

**Figure 2.**
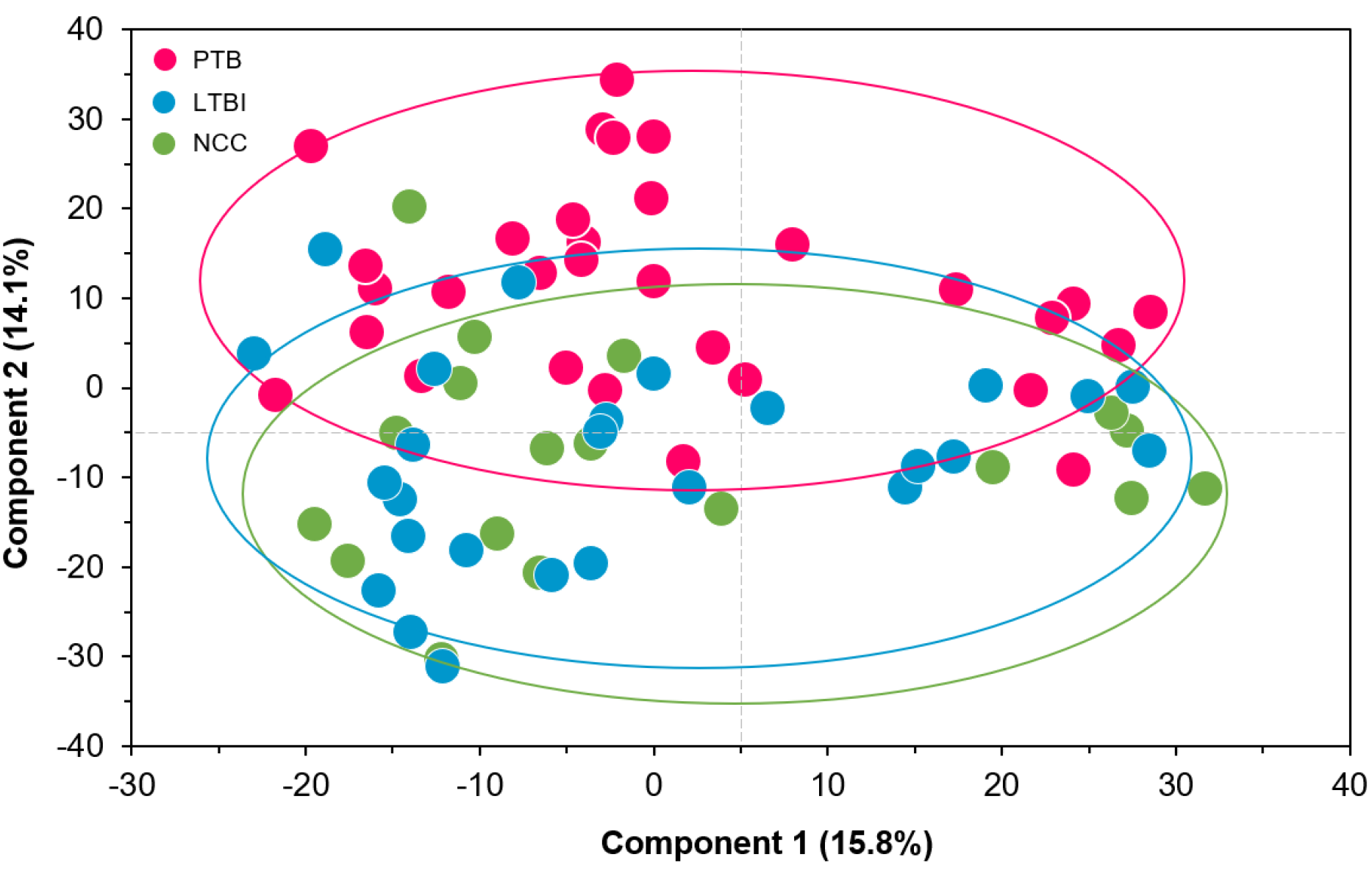
Principle component analysis (PCA) of sputum proteomes for PTB (red), LTBI (blue) and negative community controls (NCC, green). Two principle components explaining 15.8% and 14.1% of the variance among the groups are displayed. The data represent quantities of 432 human proteins. No separation of clusters is observed for the LTBI and NCC subjects while PTB datasets cluster separately.

**Table 1.** Subset of differentially abundant proteins (PTB vs LTBI).

| Protein name<sup>1</sup> | Protein SN<sup>2</sup> | Fold-change | Welch test<sup>3</sup>, -<br>log10 P-value | Extracellular or cellular<br>functions<sup>4</sup> | Body fluid and tissue<br>enrichment<sup>5</sup> |
| --- | --- | --- | --- | --- | --- |
| Ferritin | FTH1 | 5.39 | 6.60 | Acute phase response | NA |
| α-1-Acid glycoprotein 1 | ORM1 | 5.89 | 5.06 | Acute phase response | Plasma, *alveolar macs [55] |
| α-1-Acid glycoprotein 2 | ORM2 | 4.46 | 4.59 | Acute phase response | Plasma, *alveolar macs [55] |
| Neutrophil collagenase | MMP8 | 4.19 | 4.74 | ECM degradation | *Neutrophils [56] |
| Peptidoglycan recognition protein 1 | PGLYRP1 | 3.52 | 3.34 | Bactericidal protein recognizing cell wall | *Neutrophils [57] |
| C-reactive protein | CRP | 4.03 | 4.47 | Acute phase response | Plasma [58] |
| Protein S100-A7 | S100A7 | 0.10 | 7.96 | Antimicrobial, Keratinization | Esophagus, oral mucosa, tons. |
| 14-3-3 protein sigma | SFN | 0.11 | 6.59 | Keratinization, cell proliferation | Esophagus, oral mucosa, tons. |
| Cornulin | CRNN | 0.16 | 5.48 | Keratinization, Cell proliferation | Esophagus, oral mucosa, tons. |
| Aldehyde dehydrogenase 3A | ALDH3A1 | 0.26 | 4.99 | Detoxification, Cell proliferation | Esophagus, oral mucosa, tons. |
| Mucin-7 | MUC7 | 0.20 | 3.45 | Cell-protective in upper respiratory tract cells | Salivary glands [59] |
| Small proline-rich protein 3 | SPRR3 | 0.08 | 4.95 | Keratinization | Esophagus, oral mucosa, tons. |

Twelve proteins with high statistical significance comparing abundances in PTB vs LTBI proteome datasets and a fold change of >3.5 (Welch test). <sup>1</sup>common protein name; <sup>2</sup>short name as listed in UniProt database; <sup>3</sup>negative logarithmic P-value; <sup>4</sup>functions derived from UniProt database entries or literature; <sup>5</sup>enrichment in human body fluid, cell type or tissue (if no reference listed, data from Protein Atlas database [60]).

**Figure 3.**
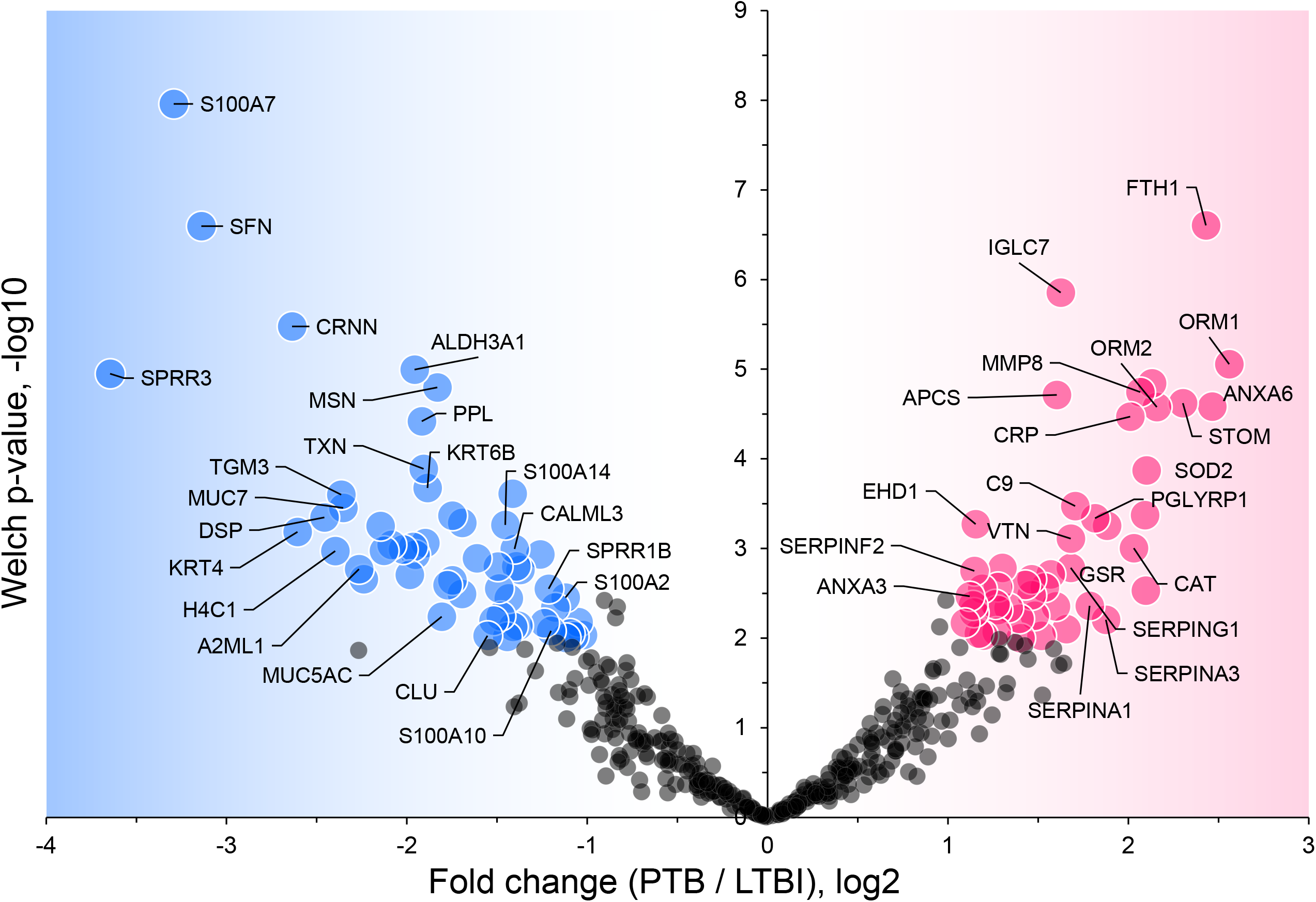
Volcano plot depicting protein abundance differences for the comparison of PTB and LTBI groups. Data are derived from sputum shotgun proteomic analyses. The unequal variance Welch t-test with multiple testing corrections was used in the Perseus software, and 103 proteins (marked with UniProt short names) had differences with a P-value < 0.01 and a fold change > 2. Red and blue dots denote proteins increased and decreased in the PTB group, respectively.

**Figure 4.**
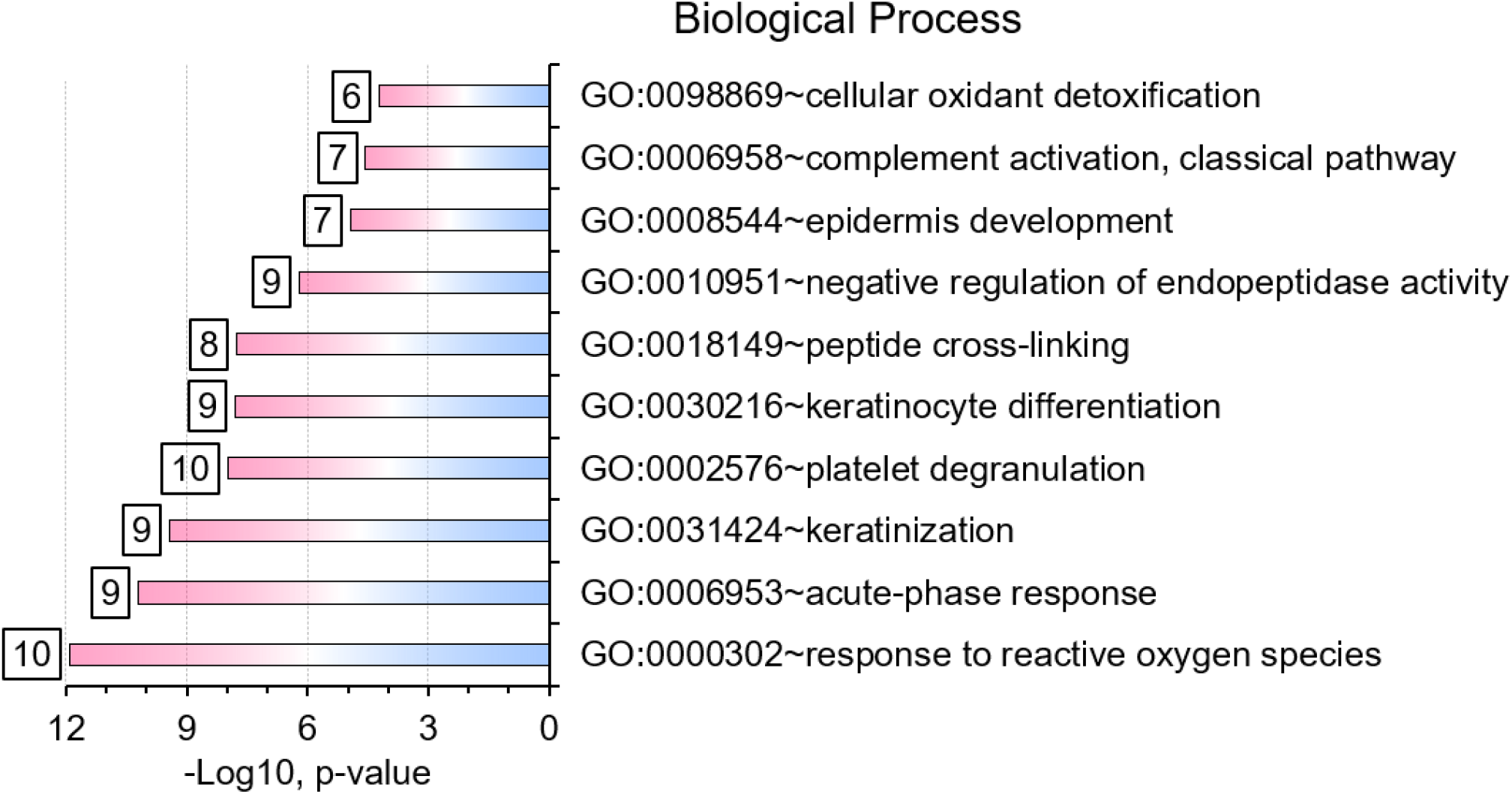
Gene Ontology (GO) biological process enrichments based on differentially abundant proteins (PTB vs. LTBI). The top-10 enriched terms and their significances (P-values) are plotted. The numbers placed next to a bar indicate the 16 proteins in our datasets belonging to that category. GO term analysis details are included in a worksheet of a Supplemental Dataset (File S5).

**Figure 5.**
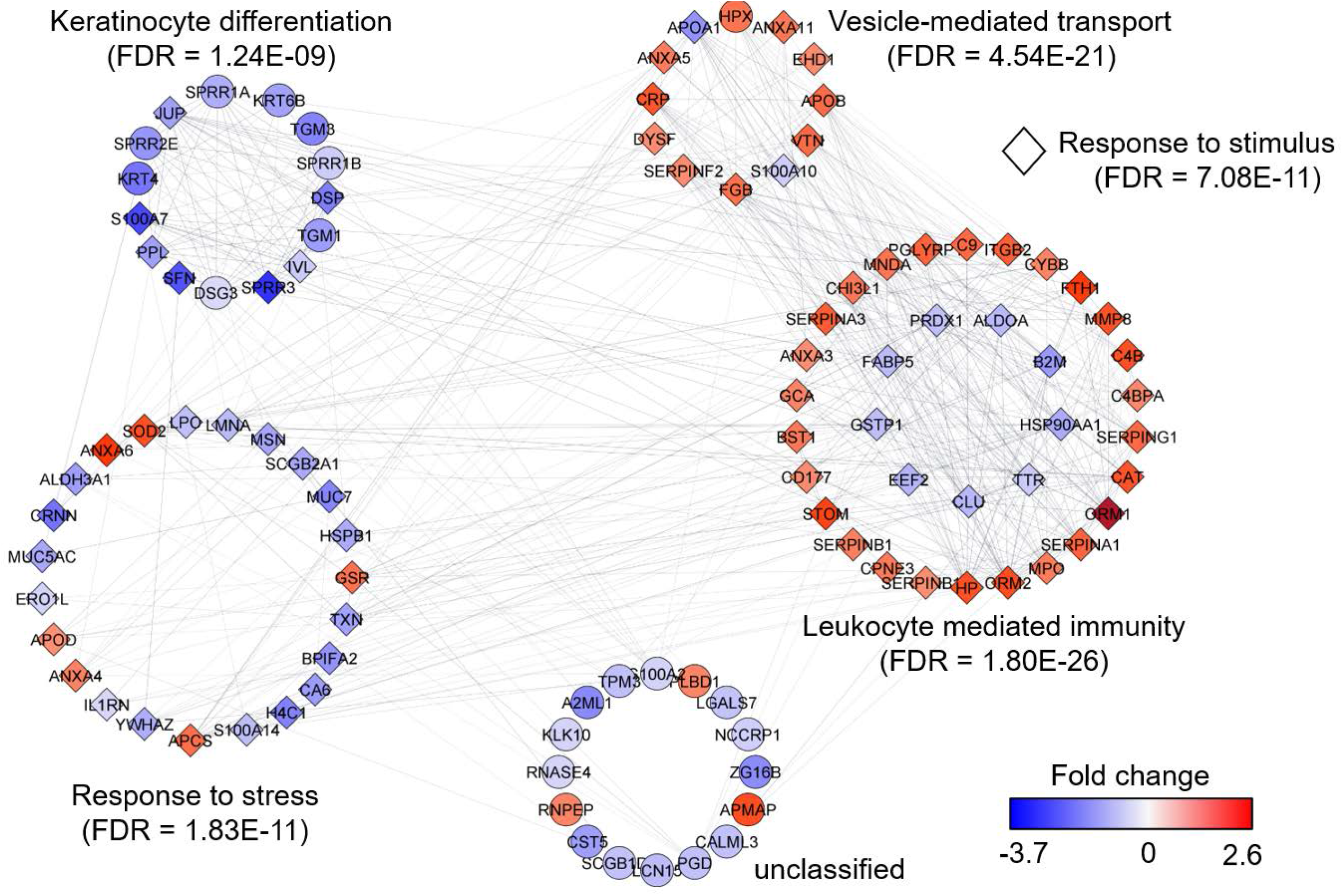
Protein network analysis and functional enrichment clusters. The network was built from 103 differentially abundant proteins comparing PTB and LTBI sample groups as input data and the String App in Cytoscape software. The score cut-off for interaction confidence was set to 0.4. Color coding is in accordance with the fold changes. Diamond shape depicts proteins associated with a response to stimulus. Protein clusters were annotated based on enrichment, a function embedded in Cytoscape.

### 3.4 α-1-Acid glycoproteins

Both complement system activation and negative regulation of endopeptidase activities are elements of the acute phase response. Proteins with these functions were increased in the PTB group (Figs. 3 and 4). In addition to ferritin, two isoforms of α-1-acid glycoprotein (α1-AGP), also named ORM1 and ORM2, had greater than 4.4-fold increases in the PTB group while no differences were measured comparing LTBI and NCC groups (Fig. 6). α1-AGP is released by alveolar macrophages during pulmonary inflammation [55], in addition to its secretion by hepatocytes into blood plasma. Western Blots for 18 sputum samples revealed a wide range of M_r_ values of α1-AGP and made proteomic data validation in a defined M_r_ range difficult (Supplementary File S6). But α1-AGP bands in PTB samples had overall higher staining intensities than LTBI and NCC samples.

**Figure 6.**
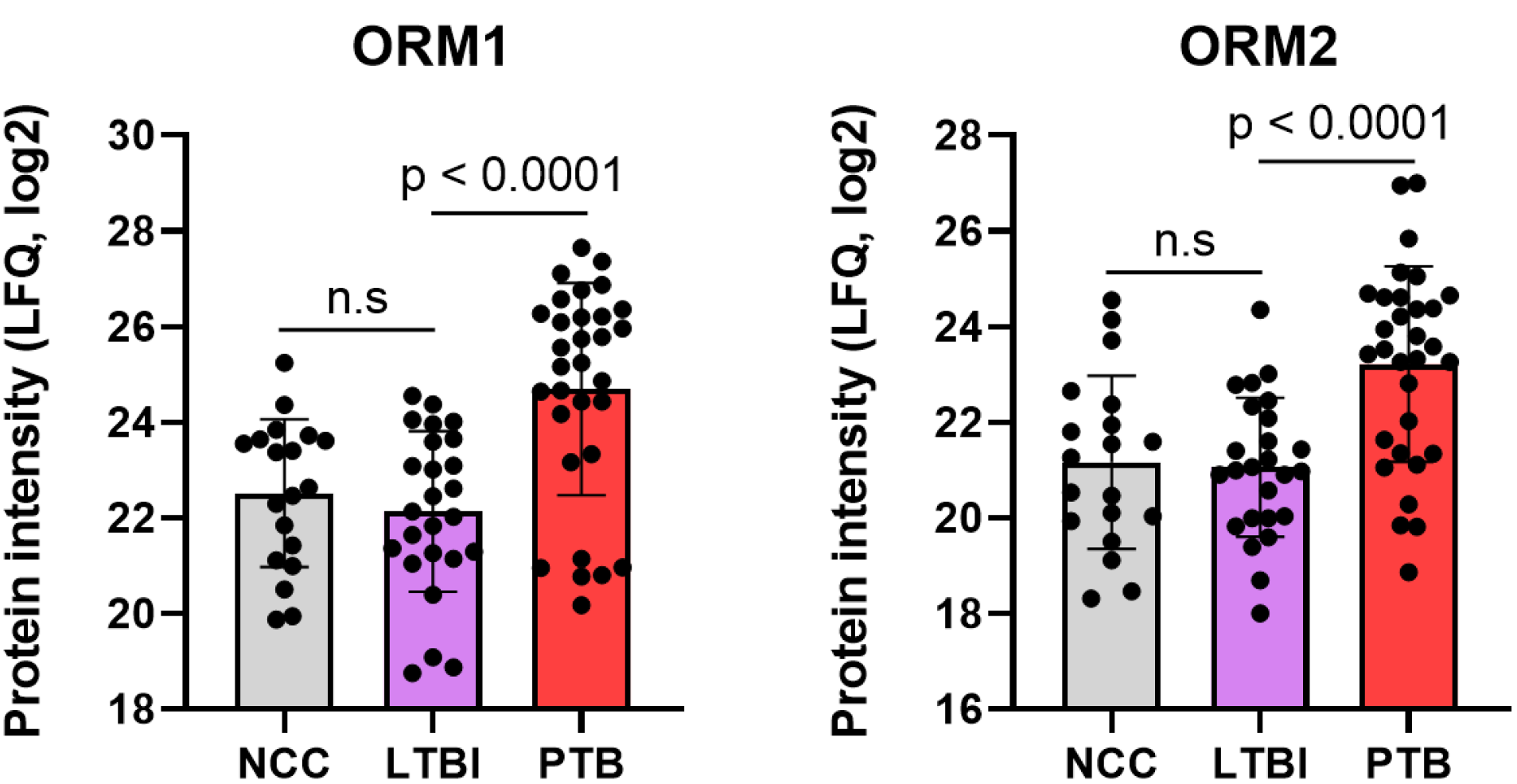
Quantitative differences for the α-1-acid glycoproteins ORM1 and ORM2 in box plots comparing datasets for PTB vs LTBI as well as PTB vs NCC. The P-values were highly significant indicating the important role for the acute phase reactants in modulating the PTB pathology. LFQ values are based on summed MS1 peak integrations for all peptides assigned to protein of origin.

### 3.5 Sputum proteomic analyses reveal a few abundance differences comparing LTBI and NCC groups

Five proteins were differentially abundant comparing the IGRA-positive and NCC datasets: lamin B1 (LMNB1), nucleobindin-1 (NUCB1), ribonuclease T2 (RNASET2), lactate dehydrogenase B chain (LDHB), and translocator protein (TSPO). Detailed data are provided in Supplementary File S5. These proteins had at least two-fold decreases for the LTBI group compared to the NCC group (Fig. 7) and may be biomarkers in sputum to discern the latent disease stage from healthy controls. Of course, if inaccuracies in IFN-γ release data and LTBI assignments occurred, differential analyses of sputum proteome data (LTBI vs NCC) are adversely affected.

**Figure 7.**
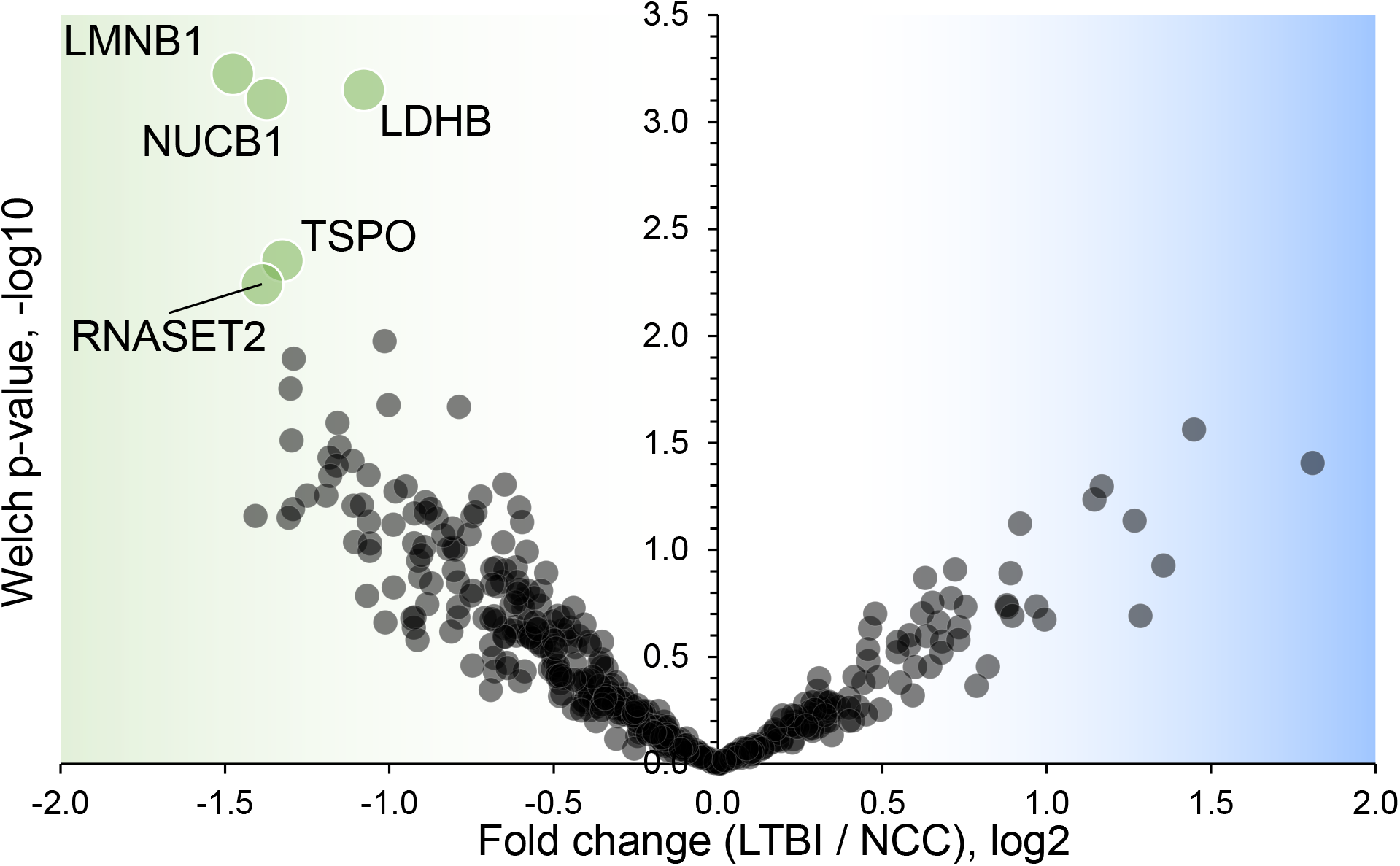
Volcano plot depicting protein abundance differences comparing the LTBI and NCC groups. A P-value < 0.01 and a fold change > 2 were applied to identify the differentially abundant proteins (each denoted in green and marked with the UniProt short name).

### 3.6 Microbiome comparisons reveal altered abundances for *Rothia* and *Haemophilus* in the comparison of PTB and LTBI cohorts

Using the V4 region of 16S rRNA to classify sputum microbial taxa, we determined *Streptococcus* to be the most abundant genus (Fig. 8) and noted the absence of α-diversity differences at the level of genera (Supplementary File S7) among the cohorts. There was no separation of the oral microbial profiles of the three cohorts based on PCA data (Supplementary File S8). *Mycobacterium*, as a genus, was detected only for individuals in the PTB group (adjusted P-value of 3 x 10^-11^, PTB vs LTBI). This finding generated confidence in the accuracy of the V4-region 16S rRNA sequence analyses (Supplementary File S9). We identified significant abundance differences for the taxa *Rothia* and *Haemophilus*. The former genus had a three-fold decrease with an adjusted P-value of 8.9 x 10^-8^; PTB vs LTBI (Fig. 9). The latter genus had a 2.2-fold increase with an adjusted P-value of 0.029; PTB vs LTBI. Interestingly, *Rothia* and *Haemophilus* were also the on average most abundant genera for their respective phyla, Actinobacteria and Proteobacteria (Fig. 8). We identified 13 additional genera with statistically significant differences (adjusted P-value < 0.05), but the fold changes were either lower than 1.5 or the genus had a low number of sequence assignments (Supplementary File S10).

**Figure 8.**
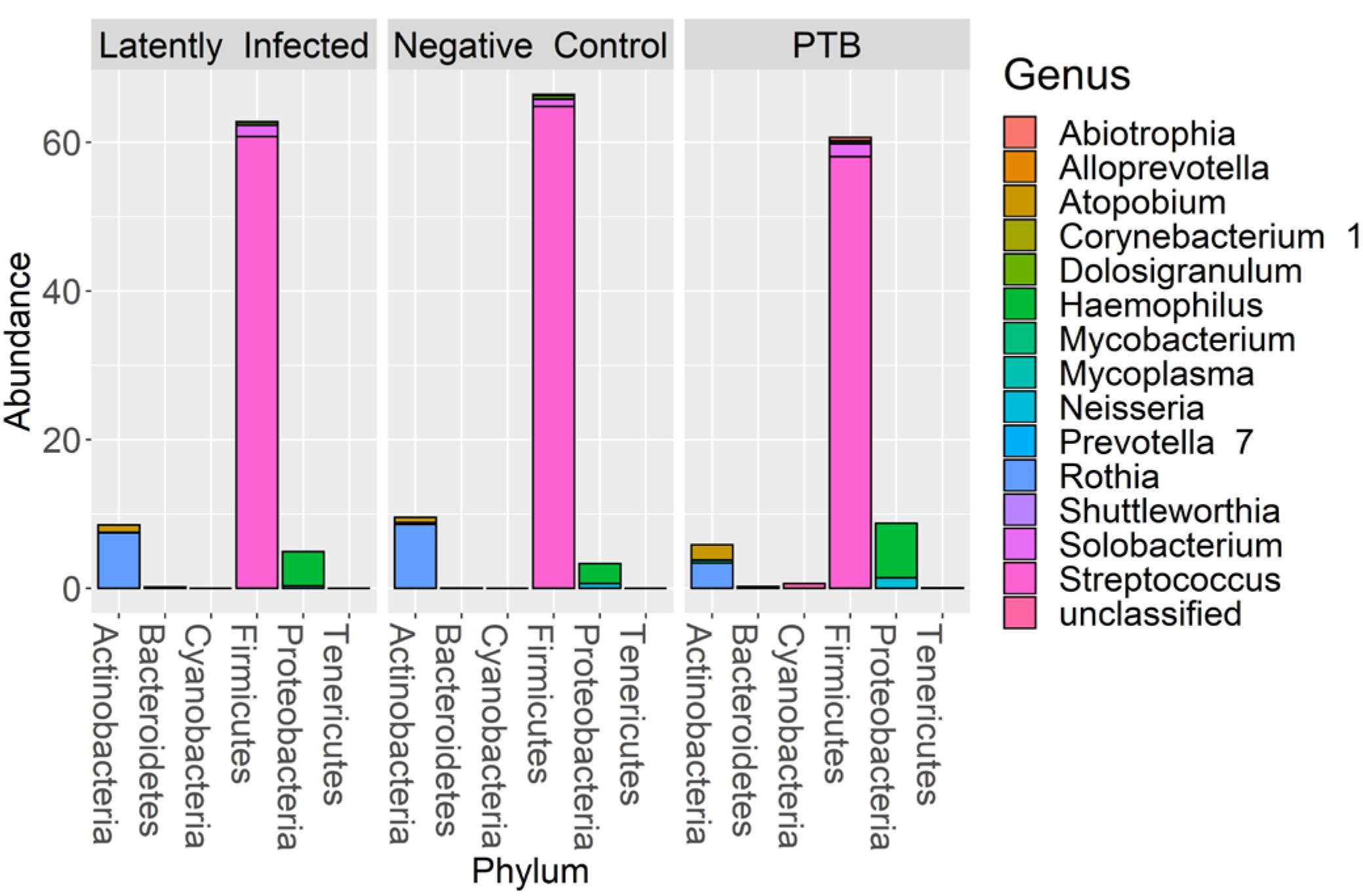
Microbial taxonomy profiling performed at the phylum level denoting the differentially abundant genera based on sequence analysis of the V4 region of 16S bacterial rRNA on a MiSeq platform. The phyla are shown denoting the most abundant genera for each phylum by color codes. Streptococcus is the most abundant genus of Firmicutes and dominant in oral microbiota. Among Actinobacteria, Rothia and Atopobium were dominant with variations in abundance among LTBI, NCC and PTB datasets. Mycobacterium, also an Actinobacterium, revealed low abundance so that it is not visualized in the segmented bars for this phylum. Haemophilus was the most abundant genus, followed by Neisseria, in the phylum Proteobacteria.

**Figure 9.**
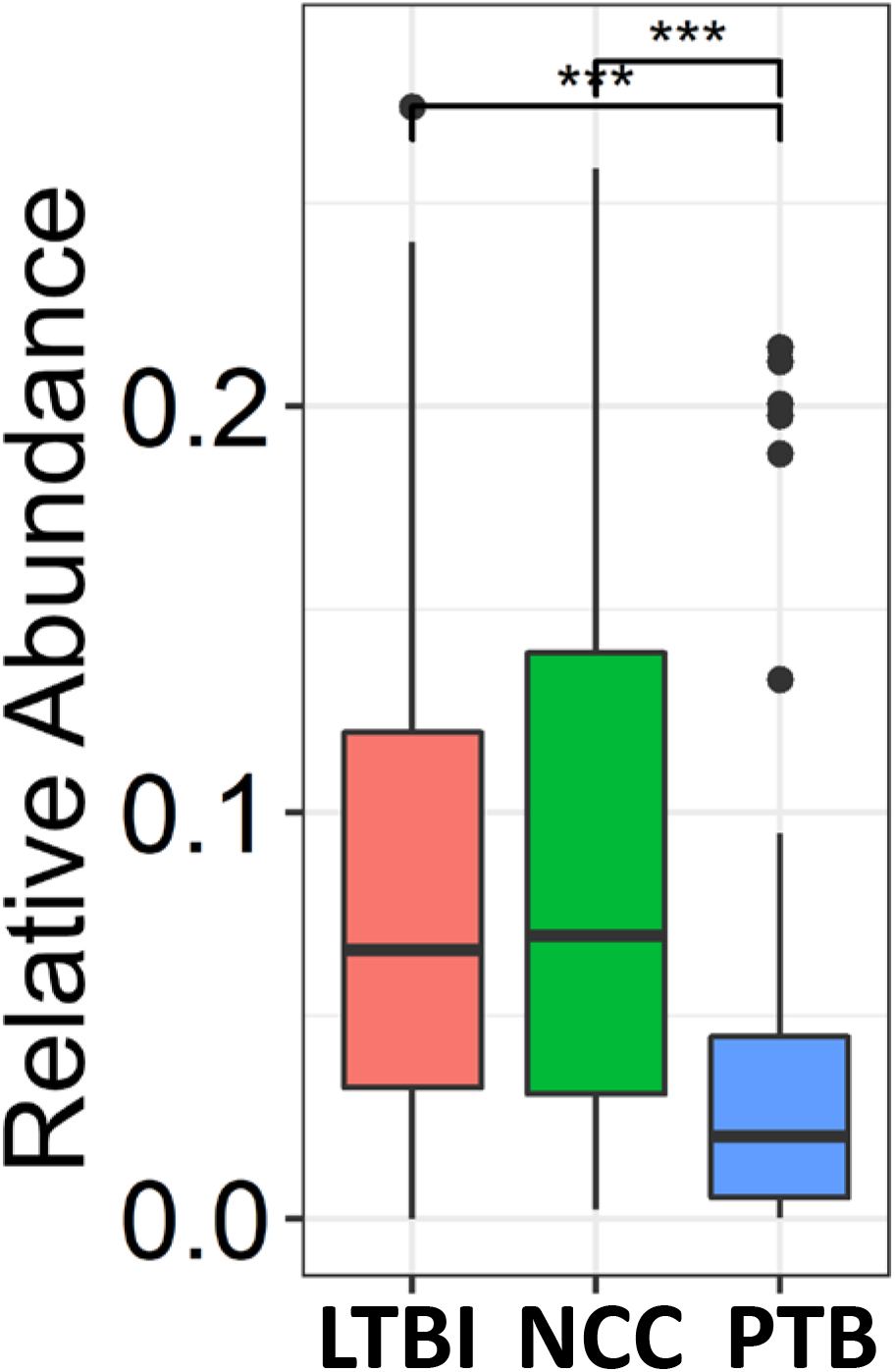
Quantitative differences for Rothia displayed in box plots comparing PTB datasets with those of LTBI and NCC groups. *** P-values were highly significant in both comparisons (< 0.001).

## 4 DISCUSSION

### 4.1 Neutrophil effectors and APR proteins are sputum biomarkers for PTB

We report quantitative differences for proteins and microbial taxa derived from sputum samples comparing a PTB cohort from South Omo with asymptomatic cohorts, one of which was designated as LTBI based on our data from a WHO-approved IGRA [14]. The study followed investigations of lineage analysis of MTBC isolates derived from individuals with TB as well as cohort and immunological characterizations of the LTBI group from this multi-ethnic pastoralist region of Ethiopia [15, 31, 32, 61]. A PCA revealed good separation of PTB proteome profiles from those of the other groups. LTBI and NCC clusters were not discernable. More than 100 differentially abundant proteins (PTB vs LTBI) allowed insights into immune responses linked to lung infection of individuals diagnosed with PTB (Figs. 3, 4, and 5). Our interpretation of functions and networks of such proteins is highly consistent with the infiltration of neutrophils (and perhaps other leukocytes) in the respiratory tract and the release of antimicrobial effectors into sputum for subjects with PTB. Examples of such effectors are peptidoglycan recognition protein 1, the collagenase MMP8, and myeloperoxidase (MPO). Proteins part of the complement system and/or APR were also increased in abundance in the PTB cohort. Other innate immune response pathways were apparently activated to eliminate the pathogen and limit or resolve host cell damage. To counteract the inflammation-generating effects of immune effectors, three serpins were elevated in abundance in the PTB group compared to the LTBI group: serpin B1, which inhibits neutrophil proteases and modulates innate immunity [62, 63]; serpin B10, which appears to control TNF-α-induced cell death [64]; and serpin G, which inhibits complement activity and other pro-inflammatory signals [65]. Oxidative stress response enzymes including catalase, SOD2, and ApoD were also increased in the PTB group, suggesting a role in controlling damage by ROS. ROS levels rise when leukocytes degranulate and activate NADPH oxidase (subunit CYBB was increased in abundance in the PTB group) and MPO. High density lipoproteins harboring ApoD and ApoB (also increased in abundance in the PTB group) are known to suppress TNF-α release from Mtb-infected macrophages [66]. Two APR proteins that scavenge heme and iron extracellularly (hemopexin and ferritin, respectively) were elevated in abundance in sputum of subjects with PTB compared to LTBI. Sequestering iron/heme limits the growth of pathogens in infected tissues [67]. Mtb is able to enter a persistent state in necrotic granulomas under iron sequestration challenges [68], thus enabling long-term survival in the host. Given that sputum proteome profiles of the LTBI group were much more similar to the NCC group than to the PTB group, the host defense and inflammation-associated pathways were apparently absent, or immeasurably small, during latent infection. The systemic role of neutrophils in the response to PTB was linked to a neutrophil-driven IFN-γ inducible transcriptional signature in whole blood that correlated with disease severity [21]. Whether persistent neutrophil activation is detrimental to clinical outcomes of chronic TB manifestations is a matter of debate [69]. Since TB severity was not assessed in our work, we were unable to correlate the APR and neutrophil biomarkers with the severity of infection.

### 4.2 Proteins of epithelial and salivary gland origin are decreased in abundance in PTB vs LTBI subjects

Most of the 56 proteins decreased in abundance in the PTB group compared to the LTBI group are enriched in squamous epithelial cells. Supported by tissue-specific expression data using the Protein Atlas Resource [60], high expression is reported for esophagus, the oral mucosa and salivary glands. Since label-free LC-MS/MS quantification calculates protein contributions relative to total proteome, we argue that decreased abundances of these proteins in sputum of PTB patients is a consequence of infiltration, degranulation and lysis of immune cells along with APR proteins that are secreted via the microvasculature into the airways. It was reported that viral respiratory infections induce mucin secretion to enable trapping the viruses in mucus [70]. We do not see an analogy to Mtb infection. Two mucins secreted into the respiratory tract (MUC7 and MUC5AC) were actually less abundant in PTB compared to LTBI sputum profiles. Psoriasin (S100-A7), a squamous epithelial protein also decreased in PTB datasets has antimicrobial and neutrophil-degranulating functions in the upper airways [71]. Thus, our data do not support a role of S100-A7 in the anti-Mtb response. We speculate that Mtb infection does not trigger host defenses in the mucosa of the upper airways, in line with the finding that upper respiratory tract TB is a rare clinical event [72].

### 4.3 α-1-Acid glycoproteins are strong sputum biomarkers for PTB

APR proteins were invariably higher in abundance in sputum of PTB compared to LTBI subjects. APR protein increases were previously linked to Mtb infections and a biomarker role: serum α-1-antitrypsin in the context of TB diagnosis [73]; serum CRP and ferritin as biomarkers of disease persistence during anti-TB therapy [18]. Ferritin concentrations were reported to be increased in acute inflammatory lung injury in order to bind extracellular and intracellular iron to reduce host susceptibility to oxidative damage [74]. Ferritin was also associated with leakage from damaged cells [75]. In addition to ferritin, we found two α1-AGP proteins (ORM1 and ORM2) to be elevated in abundance in PTB patients. Earlier, α1-AGP was described as induced in alveolar macrophages upon lung inflammation, with a 4-fold increased secretion in patients afflicted by interstitial lung injuries [55]. A murine study identified foamy macrophages located in pneumonic areas of Mtb-infected lungs as an important source of α1-AGP; administration of antibodies neutralizing this glycoprotein led to lower bacillary loads and less tissue damage, suggesting an adverse effect of α1-AGP during disease progression [76]. We surmise that sputum α1-AGPs and ferritin are surrogate biomarkers of PTB and also influence the outcome of Mtb infections. The contribution of α1-AGPs to the immunopathology of PTB needs to be evaluated further in disease models. Their roles as sensitive and specific protein biomarkers in sputum, in the context of early diagnosis or disease progression, requires further validation by surveying larger cohorts.

### 4.4 LTBI biomarker candidates

Five proteins were differentially abundant comparing LTBI and NCC cohorts (P-value < 0.01), all of them less abundant in the sputum of the LTBI group. The proteins may be biomarker candidates for LTBI diagnosis complementary to IGRAs. IGRA specificity as well as negative and positive predictive values were reported to be high for LTBI diagnosis [77]. However, the test kits are expensive and moderate performances were noted for their sensitivity (55% - 70%) to diagnose latent disease in children [78]. With respect to the proteins identified by our differential proteomic analysis, only the translocator protein TSPO was previously linked to the disease pathology and a biomarker function to monitor TB in situ [79]. Radioiodinated DPA-713, a TSPO synthetic ligand, was used to visualize anti-TB host responses in vivo. Strong TSPO and CD68 co-staining was observed for macrophages in granulomas [79]. TSPO, a protein enriched in mitochondrial outer membranes [80], is also endogenously expressed in bronchoalveolar epithelial cells [81], mediates cholesterol translocation and was physiologically linked to neuroinflammation by influencing MAPK pathways and the NLRP3 inflammasome [80]. Interestingly, Mtb activates the NLRP3 inflammasome via the ESX-1 secretion system [82]. TSPO was not differentially abundant in our PTB vs LTBI cohort comparisons. We hypothesize that low abundance of TSPO in macrophages alters the crosstalk of pathogen and immune cells in infected lungs, but this does not explain why there would be quantitative differences for this protein between LTBI and NCC cohorts. LDHB, another protein decreased in the LTBI vs NCC cohort, may also influence anti-Mtb immune responses.

The mitochondrial enzyme’s product, lactate, was reported to promote the switch of CD4 T cells to an IL-17 T cell subset and reduce CD8 T cell cytolytic capacity [83]. A third protein was ribonuclease T2, part of a family of ribonucleases that have immunomodulatory and antimicrobial properties [84].

### 4.5 Rothia, a respiratory tract microbial biomarker for PTB?

We conducted the largest microbiome profiling effort related to TB and the use of respiratory secretions to date. While there were no differences in overall α- or β-diversity in sputum microbiomes among the cohorts, we identified statistically significant differences in abundance for the genera *Rothia* and *Haemophilus*, two oral microbiome community members, in the comparison of PTB and LTBI datasets. *Rothia mucilaginosa* was recently reported to produce the siderophore enterobactin in the human oral niche [85]. Mtb is also a producer of siderophores (mycobactins) released to acquire iron from the host in infected lung tissue [86]. Enhanced iron sequestration during an active host inflammatory response in PTB possibly diminishes the fitness and growth of Rothia species in the upper respiratory tract. *Rothia* is a ubiquitous commensal organism of the oral cavity [40, 85]. One may speculate that the genus *Haemophilus* increased in abundance in respiratory secretions of PTB compared to LTBI subjects due to better adaptation to inflammatory conditions caused by infection with Mtb. The pathogen *Haemophilus* influenzae indeed adapts to the neutrophil-rich milieu in the inflamed airways of asthmatic patients [87]. Our 16S rRNA analysis did not allow species-level resolution for *Haemophilus*. Other oral microbiome studies surveying TB patient and control cohorts did not report statistically significant differences for the aforementioned genera [30]. Only one study analyzed close to 100 specimens [10] and associated sputum microbiota with different treatment outcomes for TB. LTBI subjects were not included. The most significant genus-level change was *Prevotella*, an anaerobic bacterium with higher abundance in healthy controls compared to subjects with TB [10]. A recent intriguing discovery was the role of Helicobacter colonization of the gut in IFN-γ-dependent reduced susceptibility to active TB progression, observed in a primate study that may translate to *Helicobacter* colonization in humans [88]. The production of metabolites by anaerobic respiratory bacteria was discussed as a potential modulatory influence on risk of TB progression [30]. Given the high compositional variability of oral and gut microbiota for different socio-economic and geographic settings, such comparisons remain challenging. Our study is certainly influenced by diverse diets of the many ethnic groups enrolled in the study and also the occurrence of malnutrition. Further studies will be needed to determine the functional relevance of *Rothia* and *Haemophilus* abundance changes in the context of PTB and inflammation of the human airways.

## Data Availability

The study was observational and served to understand different disease outcomes of tuberculosis via biomarker identifications. Cohort recruitment data on record are provided in Supplemental Files. Human subject enrollment and approvals were described in two previously peer-reviewed and published articles using the same cohort.
1. BMC Public Health. 2018 Feb 17;18(1):266. doi: 10.1186/s12889-018-5149-7.
2. Infect Immun. 2018 Mar 22;86(4):e00759-17. doi: 10.1128/IAI.00759-17

## 5 ACKNOWLEDGEMENTS

This work was supported by the grant 1U01HG007472 (National Institutes of Health). We appreciate the contributions of health care staff working in clinics of the South Omo province who assisted in human subject recruitment and specimen collection efforts.

## 9 SUPPLEMENTARY MATERIAL (list with headlines)

Supplementary File S1 (Table). Human subject cohort with disease category, gender, ethnic group identity from the geographic region of South Omo in Ethiopia.

Supplementary File S2 (Dataset). All protein identifications from LC-MS/MS analysis, with human and Mycobacterium tuberculosis protein IDs listed separately, and the MaxQuant data matrix for LFQ-quantified human proteins.

Supplementary File S3 (Figure). Display of correlation coefficients (R) for LC-MS/MS proteomes, technical replicates and biologically distinct samples (based on 432 quantified and quality-filtered sputum proteins).

Supplementary File S4 (Figure). Pearson Correlation hierarchical clustering applied to 75 sputum proteomic datasets. Supplementary File S5 (Dataset). Analysis of fold changes and statistical differences of proteins using MaxQuant output for 75 samples comparing PTB vs LTBI datasets with associated GO term enriched biological processes and comparing LTBI vs NCC datasets.

Supplementary File S6 (Figure). Western blots of α-1-acid glycoprotein in sputum analyzing seven TB, six LTBI and five NCC samples.

Supplementary File S7 (Figure). α-Diversity analysis of the sputum microbiomes using Chao1 and Shannon index calculations.

Supplementary File S8 (Figure). Principle component analysis of sputum microbiome datasets for pulmonary TB (PTB), latently infected, and negative community controls.

Supplementary File S9 (Figure). 16S rRNA analysis reveals presence of genus Mycobacterium in only PTB specimens supporting detectable levels of M. tuberculosis.

Supplementary File S10 (Table). Differentially abundant taxa from 16S rRNA taxonomy, OTUs and genus assignments comparing pulmonary TB (PTB) with latent infection (LTBI) specimens.

